# A brain structural connectivity biomarker for diagnosis of autism spectrum disorder in early childhood

**DOI:** 10.1101/2021.11.03.21265845

**Authors:** Xi Jiang, Xiao-Jing Shou, Zhongbo Zhao, Fanchao Meng, Jiao Le, Tianjia Song, Xinjie Xu, Xiaoyan Ke, Yuzhong Chen, Xiaoe Cai, Weihua Zhao, Juan Kou, Ran Huo, Ying Liu, Huishu Yuan, Yan Xing, Jisheng Han, Songping Han, Yun Li, Hua Lai, Lan Zhang, Meixiang Jia, Jing Liu, Keith M. Kendrick, Rong Zhang

## Abstract

**Objective:** Autism spectrum disorder (ASD) is associated with altered brain development, but it is unclear which specific structural changes may serve as potential diagnostic markers. This study aimed to identify and model brain-wide differences in structural connectivity using MRI diffusion tensor imaging (DTI) in young ASD and typically developing (TD) children (3·5-6 years old).

**Methods:** Ninety-three ASD and 26 TD children were included in a discovery dataset and 12 ASD and 9 TD children from different sites included as independent validation datasets. Brain-wide (294 regions) structural connectivity was measured using DTI (fractional anisotropy, FA) under sedation together with symptom severity and behavioral and cognitive development. A connection matrix was constructed for each child for comparisons between ASD and TD groups. Pattern classification was performed and the resulting model tested on two independent datasets.

**Results:** Thirty-three structural connections showed increased FA in ASD compared to TD children and associated with both symptom severity and general cognitive development. The majority (29/33) involved the frontal lobe and comprised five different networks with functional relevance to default mode, motor control, social recognition, language and reward. Overall, classification accuracy is very high in the discovery dataset 96.77%, and 91·67% and 88·89% in the two independent validation datasets.

**Conclusions:** Identified structural connectivity differences primarily involving the frontal cortex can very accurately distinguish individual ASD from TD children and may therefore represent a robust early brain biomarker.

## Introduction

There is an increasing consensus that children with autism spectrum disorder (ASD) have an abberant pattern of brain development.^1-2^ A number of structural magnetic resonance imaging studies using diffusion tensor imaging (DTI) have identified altered fractional anisotropy (FA), which is a widely used index in DTI to reflect axonal density and myelination, in individuals with ASD, particularly in the frontal,^3-5^ occipital lobes,^6^ and corpus callosum.^4,7^ Developmental changes in FA occur with increases in fiber tracts during the first few years, followed by decreases in later childhood and adolescence, through into adulthood.^3,5^ However, the discrimination accuracies between ASD and typically developing (TD) individuals reported by a small number of studies to date following machine learning classification approaches only found modest effects and have not been validated using independent validation datasets (75-80%^6-7^). Analysis of large fiber tracts connecting many different brain regions may also obscure changes in altered structural connectivity between specific brain regions.

In the current study we have therefore used DTI to identify differences in inter-regional structural connectivity at the whole-brain level in ASD compared to TD children in 294 different brain regions. We chose to restrict the age range of children to 3·5-6 years old since this corresponds to the period when ASD symptoms have become robustly established.^8^ Previous tractography-based research also suggests that at this age overall structural connectivity differences between ASD and TD children may be less pronounced.^4^

Based on previous studies, we firstly hypothesized that ASD children would exhibit significantly greater FA in structural connections at the whole brain level and particularly involving frontal regions compared to TD children. Secondly, we hypothesized that altered structural connections would be in networks associated with ASD symptoms and cognitive and behavioral development. Finally, we hypothesized that the structural connectivity changes identified would accurately predict ASD diagnosis at the individual level not only within the original discovery dataset but also in two independent validation datasets.

## Methods

### Participants

The present study included three independent datasets: a discovery and two validation datasets.

### Discovery dataset (Beijing)

The experiment was approved by the ethics committee of the Peking University Institutional Review Board (approval no. IRB00001052-13079). A total of 119 pre-school children either diagnosed with ASD (n = 93) or TD children (n = 26) were recruited. The age range of participants was 3·5 to 6 years, which is regarded as the time of the most severe emerging symptoms of autism.^8^ Children with ASD were recruited through pediatric psychiatric clinics and autism rehabilitation training centers in Beijing. Age and gender matched TD children were also recruited through online social platforms or day care centers in Beijing.

### ASD validation dataset (Chengdu)

The experiment was approved by the ethics committee of the University of Electronic Science and Technology of China (approval no. 1420190601). A total of 12 ASD children were recruited aged 3 to 8 years through the child healthcare department of Chengdu Women’s and Children’s Central Hospital.

### TD validation dataset (Nanjing)

The experiment was approved by the medical ethics committee of the Brain Hospital affiliated to Nanjing Medical University (approval no. KY043). A total of 9 TD children were recruited aged 4 to 6 years either through the Nanjing child mental health research center, online social platforms or day care centers.

All the participants’ parents in the three datasets were informed in detail of the research objectives and procedures, and provided written informed consents. Exclusion criteria were: (1) neurological complications, such as epilepsy, cerebral palsy, Fragile X syndrome etc.; (2) medical intervention, such as antipsychotic drugs, transcranial magnetic stimulation, acupuncture etc.; (3) diagnostic imaging anomalies or craniocerebral trauma; (4) other contraindications to MRI; (5) TD children had no family histories of any mental disorders and exhibited no evidence of developmental delay.

### Clinical Diagnosis

Participants in ASD groups were diagnosed at either Peking University Sixth Hospital or Beijing Children’s Hospital, Beijing, China for Beijing dataset, or at Chengdu Women’s and Children’s Central Hospital, Chengdu, China for Chengdu dataset. All children in the ASD group met the diagnostic criteria of Diagnostic and Statistical Manual of Mental Disorders IV-Text Revision (DSM-IV-TR)^9^ or Fifth Edition (DSM-5)^10^ and International Statistical Classification of Diseases and Related Health Problems 10^th^ revision (ICD-10)^11^. In addition, ASD diagnosis was confirmed in the Beijing dataset using the Autism Diagnostic Observation Schedule (ADOS)^12^ Traditional Mandarin version, module 1 or module 2 based on the child’s language ability. For children in the Chengdu dataset, diagnosis was confirmed using ADOS-2.^13^ Moreover, in the Beijing ASD and TD cohorts, cognitive ability was also assessed using the Gesell Developmental Scale (GDS)^14^ administered by an experienced pediatrician. This is a measure of cognitive and behavioral development and adaptability including five components (gross motor, fine motor, adaptive, language and personal social behaviors).

### MRI Acquisition and Preprocessing

Children in both ASD and TD groups of the three datasets were sedated by oral administration of chloral hydrate at the 50 mg/kg body weight (1 g maximum dose), commonly used for pediatric clinical imaging. During the MRI scan, children wore earplugs and de-noising headsets to reduce the noise, and parents were encouraged to remain in the scanning room to ensure safety in case the child awoke. In all sites, brain images were reviewed by neuroradiologists to confirm absence of neurological abnormalities.

For the discovery dataset MRI images were acquired on a GE 3T MR750 scanner with a 12-channel head coil at the Peking University Third Hospital. DTI data were obtained with an echo-planar imaging sequence: TR = 9,000 ms, TE = 89·4 ms, FOV = 256 mm, matrix size = 128 × 128, voxel size = 2 mm isotropic, 75 slices covering the whole brain with no gap, 32 diffusion directions, b-value = 1000 s/mm^2^. For independent ASD and TD validation datasets see supplementary methods. Pre-processing methods for DTI raw data are described in the supplementary methods.

### Overview of analysis

The flow chart of ASD identification framework is illustrated in Fig. 1. Based on the pre-processed DTI FA map (Fig. 1(A)), the whole-brain streamline fibers were reconstructed (Fig. 1(B)) and adopted to the brain atlas (Fig. 1(C)). To construct the structural connection matrix for each subject (Fig. 1(D)), we calculated the pair-wise FA value between every two brain regions. The constructed structural connection matrix was further projected into the brain for each participant (Fig. 1(E)). Group comparisons were then performed based on all structural connection networks between ASD and TD groups (Fig. 1(F)) to obtain the between-group differences (Fig. 1(G)). The resulting connections were finally adopted as features to perform pattern classification between ASD and TD groups (Fig. 1(H)). The details of each step are demonstrated in the following sections.

**Figure 1.**
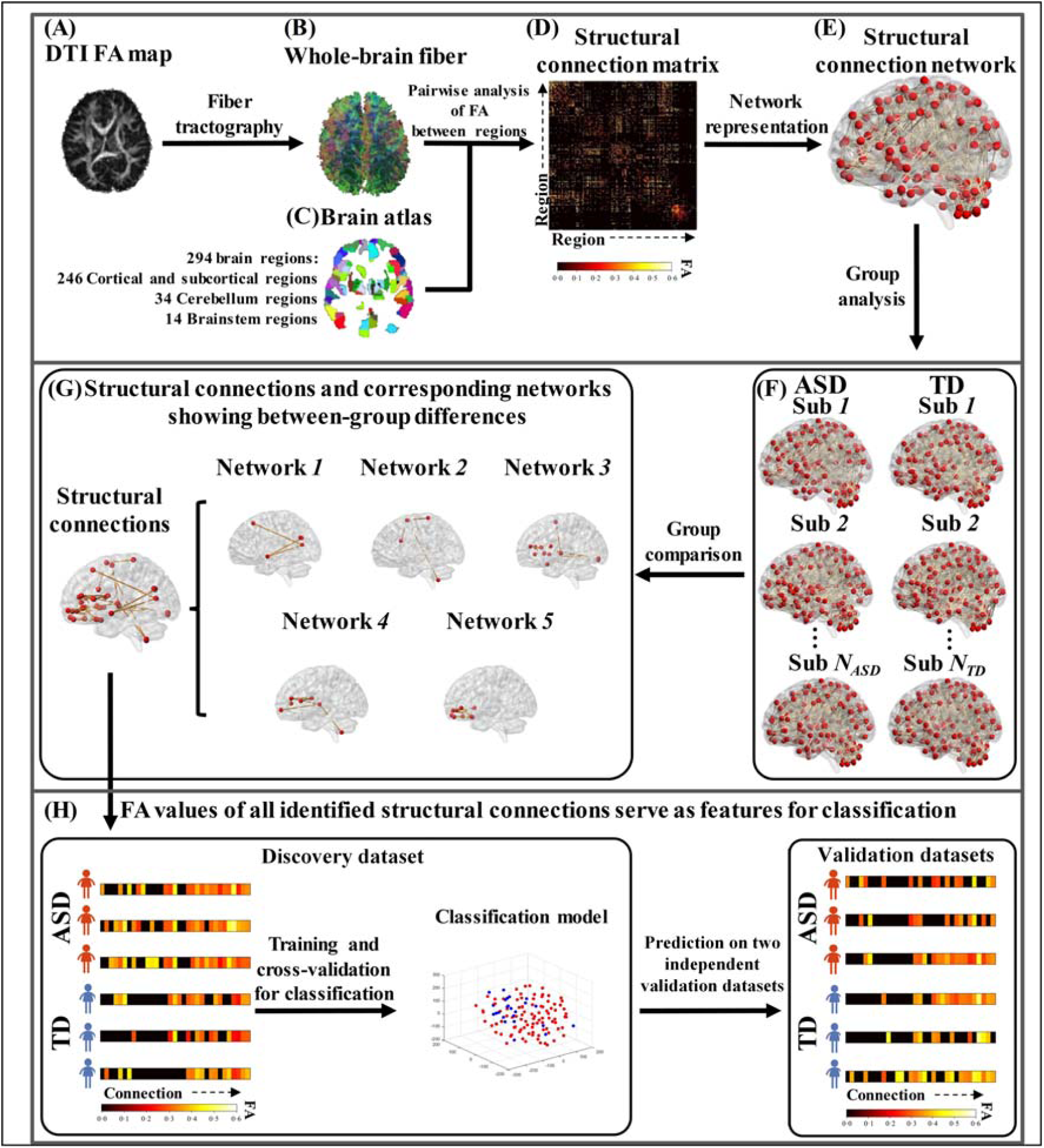
Flow chart of ASD identification framework. **(A)** The pre-processed DTI FA map. **(B)** Reconstructed whole-brain streamline fibers. **(C)** Young children’s brain atlas. **(D)** Structural connection matrix for each participant. **(E)** The individual structural connection network. **(F)** All individual structural connection networks in ASD and TD groups. **(G)** The structural connections and associated networks showing differences between ASD and TD groups. **(H)** Pattern classification between ASD and TD using all structural connections showing between-group differences.

### Construction of Structural Connection Network

The brain atlas used in this study included 294 non-overlapping brain regions consisting of 246 cortical and subcortical regions, 34 cerebellar regions, and 14 brainstem regions (Fig. 1(C)). The cortical and subcortical regions were from the Brainnetome Atlas^15^ with 123 homotopic regions in each hemisphere; cerebellar regions were from the Human Cerebellar Probabilistic Magnetic Resonance Atlas^16^ and brainstem regions were from the Human Brainstem Standard Neuroanatomy Atlas^17^. Since the three atlases were originally defined in the adult MNI152 standard space, we first performed linear registration to warp the T1-weighted adult MNI152 image to the space of T1-weighted template image of children aged 4·5-8·5 years (https://www.mcgill.ca/bic/software/tools-data-analysis/anatomical-mri/atlases/nihpd),^18^ and then applied the linear transformation to the brain atlas to warp it to the children’s template image space as well in order to obtain the young children’s brain atlas.

The reconstructed whole-brain streamline fibers in each child (Fig. 1(B)) were then aligned to the young children’s brain atlas space via DSI Studio^19^. The structural connection matrix for each child (Fig. 1(D)) was obtained by calculating the pair-wise mean FA value between every two brain regions via DSI Studio^19^, and further represented as a structural connection network (Fig. 1(E)) in which the nodes were the brain regions and edges were the mean FA values between two nodes.

### Identification of Structural Connections Showing Differences between ASD and TD

Based on the structural connection networks of all children in the ASD and TD groups (Fig. 1(F)), we adopted the widely used Network-based Statistic (NBS) approach^20^ to identify the structural connections and associated networks showing between-group differences (Fig. 1(G)). As a non-parametric statistical method, NBS first performs a large number of univariate hypothesis tests on all edges in the network, then clustering-based statistics, and finally permutation tests to calculate the family-wise error rate (FWER) corrected *p*-values for each sub-network consisting of edges with group differences. We adopted the NBS Connectome toolbox implemented in Matlab^20^ to perform the analysis. The structural connection networks of all participants in ASD and TD groups were the inputs with gender and age as covariates. Next we performed the NBS analysis to identify both increased and decreased FA values of structural connections and associated networks in ASD compared to TD.

### Pattern Classification between ASD and TD Children

Based on the identified structural connections showing between-group differences (Fig. 1(G)), we further adopted FA values of those connections as features to perform pattern classification between the ASD and TD groups. We used the discovery dataset as the training dataset to establish the classification model. For training and leave-one-out cross-validation, we employed the widely used support vector machine (SVM) approach. The training model was then applied to the two independent validation datasets to validate its generalizability. Specifically, we adopted the widely used cost-support vector classification (C-SVC), and the radial basis function (RBF) as the kernel function in SVM. The optimal values of parameters c (i.e., cost, c ranged from 15 to 16) and g (i.e., gamma, g ranged from 0·08 to 1) in RBF were obtained by a hyperparameter optimization framework of optuna.^21^

### Potential Effect of Imbalanced Sample Size between ASD and TD

In view of the imbalanced group sample sizes in the discovery dataset (93 ASD and 26 TD), we adopted both up-sampling and down-sampling approaches to alleviate the potential model overfitting as well as classification bias problem (see supplementary methods). Both approaches confirmed that our classification model was not influenced by the imbalanced sample sizes.

### Statistical Analysis

Independent two-sample *t*-tests were utilized to identify the different structural connections between ASD and TD (n=10,000 permutation times, significance level *p*<0·05, FWE corrected) in NBS. Independent sample *t*-tests were used for continuous variables including age, BMI, head circumference, GDS total score, and FA between ASD and TD. Chi-square tests were used for categorical variables including gender and handedness between ASD and TD. Pearson’s linear correlation coefficients were computed between the averaged FA value and ADOS and GDS scores (one-sample *t*-tests, significance level at *p*<0·05, FDR corrected). A mediation model was conducted (PROCESS)^22^ using bootstrap analysis to investigate the relationship between the averaged FA value, ADOS total and GDS total scores (bootstrap = 1000).

## Results

### Subject Demographics and Behavioral Measures

Table 1 summarizes demographic and other information for ASD and TD groups in the different datasets and ADOS scores for ASD children. There were no group differences in age, gender, BMI, handedness, and head circumference in the discovery dataset, although as expected the total GDS score was significantly less in the ASD group indicating impaired cognitive and behavioral development.

**Table 1.**
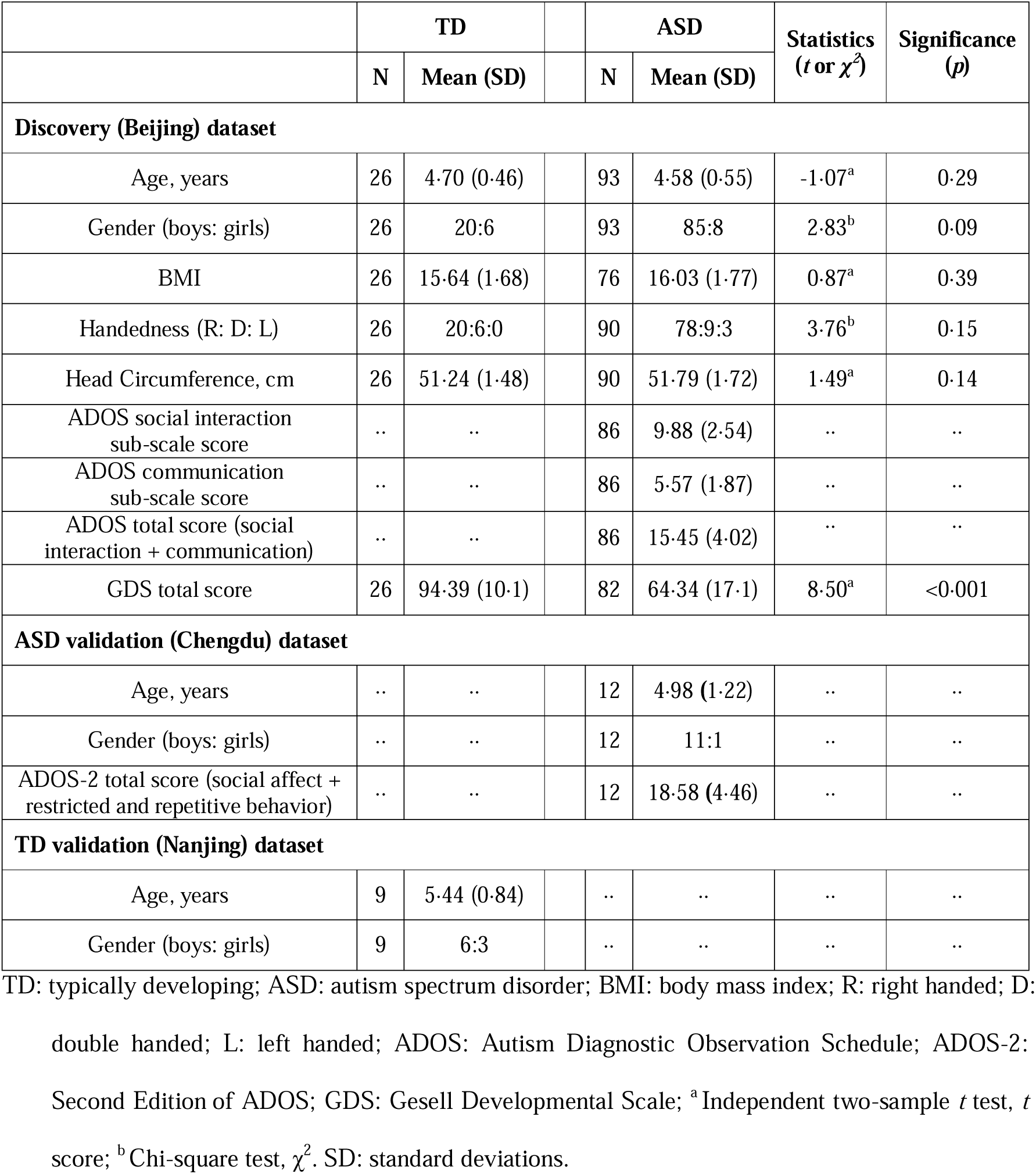
Subject demographics and behavioral measures of the three datasets.

### Increased FA Connections and Networks in ASD Compared to TD Children

We identified 33 increased but no decreased FA values of structural connections in ASD compared to TD children (Figs. 2(A) and 2(C), Table 2). Notably, 29 out of 33 connections were associated with the frontal lobe. Moreover, 30 out of 33 connections were intra-hemispheric. Fig. 2(B) illustrates the locations of 33 connections on the cortical surface. In the ASD group averaged FA values were significantly negatively correlated (FDR corrected) with ADOS total and ‘social interaction’ sub-scale scores (Fig. 2(D)) and positively correlated with the GDS total score (Fig. 2(E)). In the TD group there was a slight but not significant negative correlation between averaged FA values and GDS score (Fig. 2(E)). A mediation analysis indicated that within the ASD group the ADOS total score was the main mediator of the effects on the averaged FA value and GDS total score (Fig. 2(F)).

**Figure 2.**
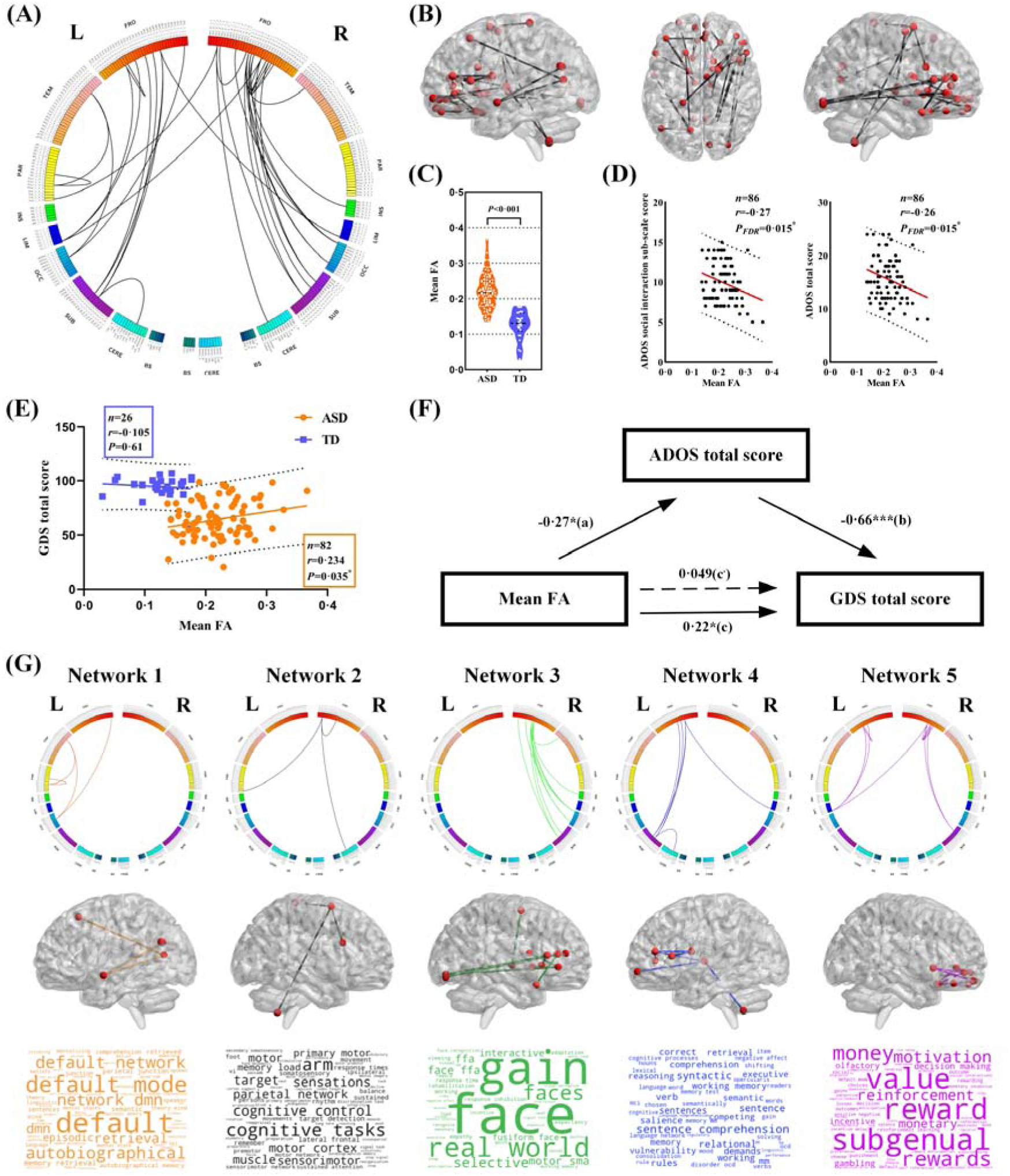
Increased FA connections and networks in ASD compared to TD children. **(A)** The 33 increased FA values of structural connections in ASD in circos plot. Abbreviations: L, left hemisphere; R, right hemisphere; FRO, frontal lobe; TEM, temporal lobe; PAR, parietal lobe; INS, insular lobe; LIM, limbic lobe; OCC, occipital lobe; SUB, subcortical nuclei; CERE, cerebellum; BS, brain stem. **(B)** Locations of 33 connections on the cortical surface. **(C)** Averaged FA value of 33 connections in ASD and TD groups. **(D)** Correlations between averaged FA value of the 33 connections in ASD and ADOS ‘social interaction’ sub-scale score and total (‘communication’ + ‘social interaction’) score. **(E)** Correlations between averaged FA value of the 33 connections and ADOS social communication and total scores and GDS total score. **(F)** Mediation analysis between averaged FA, GDS total score and ADOS total score (path a=-0·27, *p*=0·018; path b=-0·66, *p*<0·001; path c=0·22, *p*=0·047; path c, =0·049, *p*=0·58). **(G)** The 33 increased connections were further categorized into 5 structural networks. In each network, row 1-3 shows the structural connections in circos plot, locations of the connections on cortical surface, and the word cloud of functional annotation via meta-analysis respectively.

**Table 2.** Profiles of the 33 structural connections with increased FA in children with ASD.

| No. | Network | Region name | MNI coordinates |  |  | Region name | MNI coordinates |  |  |
| --- | --- | --- | --- | --- | --- | --- | --- | --- | --- |
|  |  |  | X | Y | Z |  | X | Y | Z |
| 1 | 1 | STG_L | -55 | -3 | -10 | PCun_L | -12 | -67 | 25 |
| 2 |  | IPL_L | -47 | -65 | 26 | PCun_L | -12 | -67 | 25 |
| 3 |  | SFG_L | -18 | 24 | 53 | MVOcC_L | -13 | -68 | 12 |
| 4 |  | STG_L | -55 | -3 | -10 | MVOcC_L | -13 | -68 | 12 |
| 5 | 2 | SFG_R | 20 | 4 | 64 | IFG_R | 45 | 16 | 25 |
| 6 |  | SFG_R | 20 | 4 | 64 | PoG_L | -21 | -35 | 68 |
| 7 |  | SFG_R | 20 | 4 | 64 | Right_IX | 6 | -54 | -50 |
| 8 | 3 | IFG_R | 48 | 35 | 13 | LOcC_R | 32 | -85 | -12 |
| 9 |  | IFG_R | 54 | 24 | 12 | LOcC_R | 32 | -85 | -12 |
| 10 |  | MFG_R | 42 | 44 | 14 | MVOcC_R | 10 | -85 | -9 |
| 11 |  | IFG_R | 48 | 35 | 13 | MVOcC_R | 10 | -85 | -9 |
| 12 |  | IFG_R | 51 | 36 | -1 | MVOcC_R | 10 | -85 | -9 |
| 13 |  | IFG_R | 51 | 36 | -1 | BG_R | 22 | 8 | -1 |
| 14 |  | IFG_R | 51 | 36 | -1 | BG_R | 14 | 5 | 14 |
| 15 |  | IFG_R | 48 | 35 | 13 | STG_R | 47 | 12 | -20 |
| 16 |  | IFG_R | 54 | 24 | 12 | INS_R | 36 | 18 | 1 |
| 17 |  | SFG_R | 7 | -4 | 60 | Tha_R | 12 | -14 | 1 |
| 18 |  | IFG_R | 48 | 35 | 13 | Tha_R | 12 | -14 | 1 |
| 19 | 4 | MFG_L | -41 | 41 | 16 | BG_L | -14 | 2 | 16 |
| 20 |  | MFG_L | -41 | 41 | 16 | Tha_L | -7 | -12 | 5 |
| 21 |  | IFG_L | -53 | 23 | 11 | BG_L | -14 | 2 | 16 |
| 22 |  | MFG_L | -41 | 41 | 16 | CG_R | 5 | 41 | 6 |
| 23 |  | MFG_L | -26 | 60 | -6 | Tha_L | -7 | -12 | 5 |
| 24 |  | Tha_L | -7 | -12 | 5 | Left_IX | -8 | -54 | -48 |
| 25 | 5 | OrG_R | 23 | 36 | -18 | OrG_R | 6 | 57 | -16 |
| 26 |  | OrG_R | 23 | 36 | -18 | OrG_R | 9 | 20 | -19 |
| 27 |  | OrG_L | -6 | 52 | -19 | OrG_L | -10 | 18 | -19 |
| 28 |  | OrG_R | 6 | 47 | -7 | OrG_R | 9 | 20 | -19 |
| 29 |  | OrG_L | -7 | 54 | -7 | CG_L | -4 | 39 | -2 |
| 30 |  | OrG_R | 6 | 47 | -7 | CG_L | -4 | 39 | -2 |
| 31 |  | OrG_L | -10 | 18 | -19 | CG_L | -4 | 39 | -2 |
| 32 |  | OrG_R | 6 | 47 | -7 | BG_R | 15 | 14 | -2 |
| 33 |  | OrG_R | 6 | 57 | -16 | BG_R | 15 | 14 | -2 |

Abbreviations: L, left; R, right; STG, superior temporal gyrus; PCun, precuneus; IPL, inferior parietal lobule; SFG, superior frontal gyrus; MVOcC, medioventral occipital cortex; IFG, inferior frontal gyrus; PoG, postcentral gyrus; LOcC, lateral occipital cortex; Tha, thalamus; MFG, middle frontal gyrus; OrG, orbitofrontal gyrus; CG, cingulate gyrus; BG, basal ganglia; INS, insula.

Connections with increased FA were further categorized into 5 structural networks via NBS (Fig. 2(G) and Table 2) together with a functional characterization using the Neurosynth platform, and visualized as a word cloud. Network 1: default mode function and memory retrieval, including: left superior temporal gyrus, precuneus, inferior parietal lobule, superior frontal gyrus, and medioventral occipital cortex; Network 2: motor function, including: right superior and inferior frontal gyri, right cerebellum lobe IX and left postcentral gyrus; Network 3 visual and facial recognition function, including: right inferior and superior frontal gyri, lateral occipital cortex, pre-motor thalamus, middle frontal gyrus, medioventral occipital cortex, basal ganglia, superior temporal gyrus and insula; Network 4 language and cognitive function, including: left middle and inferior frontal gyri, thalamus, cingulate gyrus, basal ganglia and cerebellum lobe IX; Network 5: social and general reward functions, including: bilateral orbitofrontal and cingulate gyri and basal ganglia.

### Classification Accuracy between ASD and TD Children

We first up-sampled the discovery dataset to 186 subjects with 93 ASD and 93 TD and trained the classification model in a 33-dimensional feature space based on the 33 connections using a leave-one-out cross-validation strategy. Fig. 3(A) shows the classification model in a 3-dimensional feature space after performing dimensionality reduction using the t-distributed stochastic neighbor embedding (t-SNE) algorithm.^23^ The Receiver Operating Characteristic (ROC) curve of the training model is shown in Fig. 3(B). The area under the ROC curve (AUC) was 0·981, indicating the robustness of the training model. The confusion matrix of the training model is shown in Fig. 3(C). Accuracy, sensitivity, specificity, precision, and F measure in both discovery and validation datasets are reported in Fig. 3(D) with the proposed model achieving high classification accuracy in both discovery (96·77%) and independent validation datasets (91·67% and 88·89%). The alternative down-sampling strategy of the discovery dataset by 1,000 times also achieved high classification accuracy in both discovery (94·85±1·30%) and independent validation datasets (91·63±5·55% and 80·04±5·52%). Thus overall, the classification model showed both high accuracy and generalizability for ASD identification across different datasets without being influenced by the imbalanced sample sizes of the discovery dataset.

**Figure 3.**
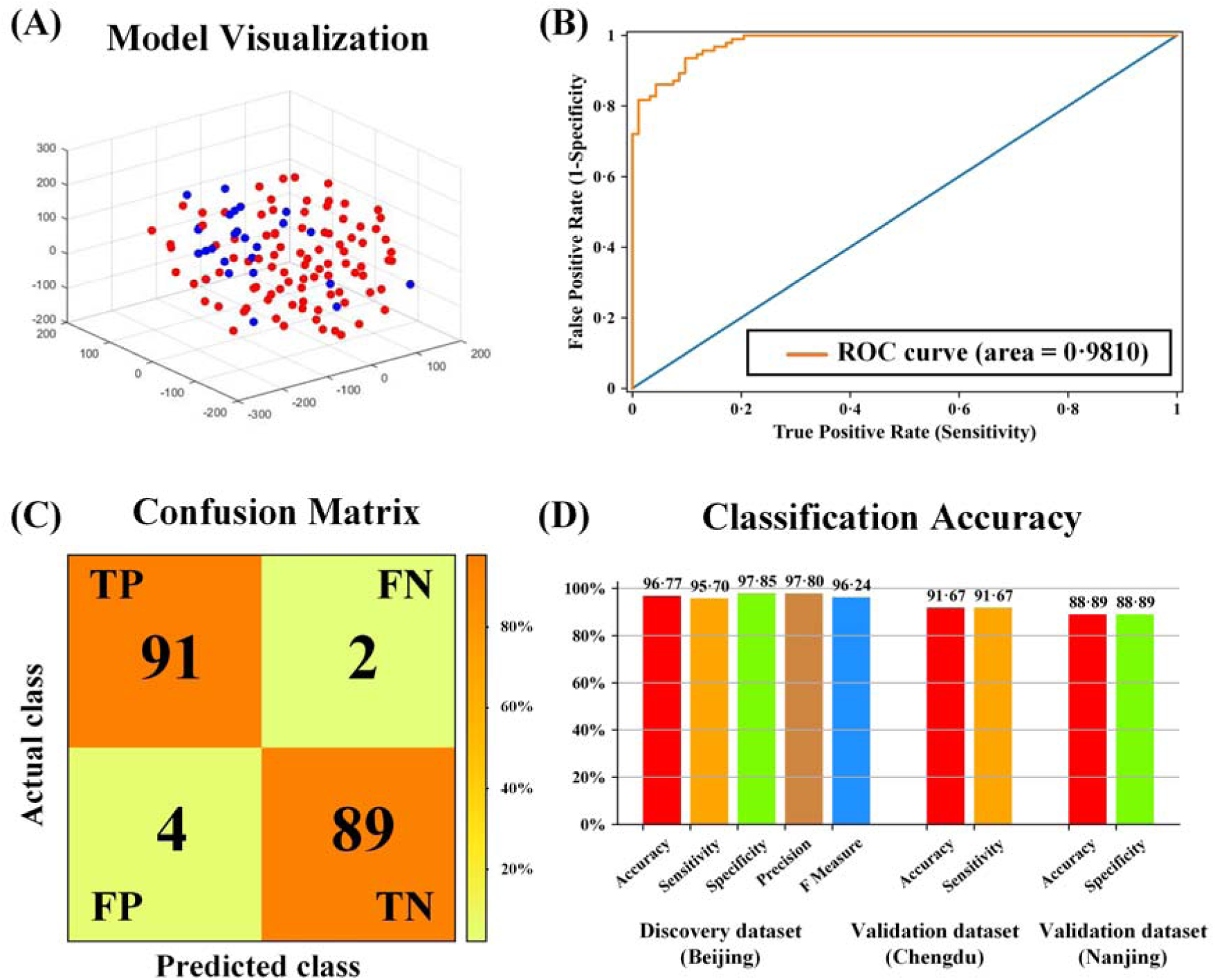
Classification Accuracy between ASD and TD. **(A)** Classification model in a 3-dimensional feature space after performing dimensionality reduction using the t-distributed stochastic neighbor embedding (t-SNE) algorithm. Red and blue dots represent ASD and TD subjects, respectively. **(B)** The Receiver Operating Characteristic (ROC) curve of the training model. **(C)** The confusion matrix of the training model. The colorbar represents the proportion of correctly classified subjects among all subjects. **(D)** The detailed classification accuracy metrics including accuracy, sensitivity, specificity, precision, and F measure in both discovery and validation datasets.

## Discussion

Our findings have revealed the presence of a small number of inter-regional structural connections within the brains of young children with ASD which exhibit increased FA compared to TD and negatively associated with symptom severity. The majority of affected regional connections involve the frontal cortex and overall they achieved a classification accuracy of 96·77% for discriminating between ASD and TD individuals in the discovery dataset and 91·67% and 88·89% in two small independent datasets. The 33 inter-regional structural connections could be clustered into 5 independent networks with relevance to a range of behavioral functions influenced in ASD.

In support of our original hypothesis, the majority of the 33 structural connections showing increased FA in children with ASD in the current study involved the frontal lobe including intrinsic short range frontal-frontal connections and longer range frontal-occipital, frontal-thalamic and frontal-limbic ones. This is in agreement with findings from other studies^3-4,24-26^ and supports the conclusion that alterations in both intrinsic and extrinsic frontal lobe structural connectivity contribute fundamentally to ASD.

The altered structural connections in children with ASD could be clustered into 5 individual networks encompassing default mode, motor, visual and facial recognition, language and memory and reward functions. The largest single frontal lobe cluster involved orbitofrontal regions and their connections with the basal ganglia. These intrinsic frontal connections are strongly associated with social and other types of reward processing as well as decision making^27^ and these functions are known to be impaired in ASD.^28-29^ Three other clusters involved inferior, medial and superior frontal gyri connections with thalamus, basal ganglia, cingulate, insula, occipital cortex, post-central gyrus and cerebellum associated with social cognition, language comprehension, sensory and motor processing functions,^30-32^ all of which are impaired in ASD.^33-36^ The remaining cluster primarily involved connections between the superior temporal gyrus and inferior parietal lobule with the precuneus in the default mode network, associated with self-processing, experience of agency, autobiographic and episodic memory retrieval and visuospatial imagery. Default mode dysfunction has been consistently reported in ASD^37^ as well as impaired self-processing, sense of agency, autobiographical and episodic memory.^38-39^

A previous study using DTI measures and classification techniques to identify ASD compared to TD children employed shape representations of white matter fiber tracts as features, and achieved 75·34% discrimination accuracy using a leave-one-out cross-validation approach.^7^ Another study adopted the anisotropy scores of regions of interest as features, and achieved 80% accuracy using leave-one-out cross-validation.^6^ In our current study, we adopted the DTI-derived FA values of structural connections as features, and achieved a much higher classification accuracy in both the discovery dataset (96·77%) and, importantly, in independent validation datasets (91·67% and 88·89%), demonstrating satisfying classification and generalization ability of our model across different datasets.

Unexpectedly, we found a significant negative correlation between the averaged FA values of the 33 altered connections in children with ASD and ADOS total and social sub-scale scores, indicating that symptom severity was actually lower in children with greater FA. Scores on GDS were also positively correlated with FA values in the ASD group but slightly negatively correlated in the TD group. A mediation analysis identified that ADOS scores were primarily mediating both FA values and GDS scores in the ASD group. This may indicate an experience-dependent compensatory effect is occurring in children with ASD whereby increased FA contributes to reduced symptom severity and enhanced cognitive and behavioral development. A social experience compensation effect has previously been described in behavioral studies of autism.^40^ Interestingly, a tractography study has reported a positive association between increased frontal lobe FA and symptom severity in very young children but a negative one in older children in the age-range of the current study.^4^ Thus, children who experience more severe symptoms at the age of 3.5-6 years may have reduced FA in these neural circuits compared to when they were younger, whereas those with milder symptoms may instead have maintained or even increased their FA. A longitudinal study would clearly be required to confirm this possibility.

## Limitations

A limitation of the current study is its cross-sectional nature and restricted age range (3·5-6 years old). Patterns of structural differences may differ in both younger and older individuals and only a longitudinal design study can address this. A second limitation is we did not determine whether observed changes are specific to ASD or might also occur in children with developmental delay, for example. A final limitation is that we could only obtain two small datasets for independent analysis of discrimination accuracy although the findings were very encouraging.

## Conclusion

By employing a fine-grained, brain-wide analysis of structural differences for regional connections in the brains of young (3·5-6 years old) autistic compared with typically developing children we have identified a number of structural connections mainly involving the frontal lobe exhibiting increased FA but negatively associated with symptom severity. Differences in these structural connections show high accuracy (>96%) in discriminating autistic children from TD children which generalizes to independent novel datasets. These new findings suggest that differences in structural connections primarily involving the frontal cortex of young autistic children are a potential reliable and generalizable biomarker for ASD diagnosis and for assessing the efficacy of therapeutic interventions.

## Data Availability

All data produced in the present study are available upon reasonable request to the authors

## Contributors

XJ: conceptualization, methodology, software, validation, formal analysis, resources, data curation, writing - original draft, writing - review & editing, visualization, supervision, project administration, funding acquisition. XJS: methodology, validation, investigation, resources, data curation, writing – review & editing, project administration, funding acquisition. ZBZ: methodology, software, validation, formal analysis, visualization. FCM: investigation. JL: investigation. TJS: investigation. XJX: investigation. XYK: resources. YZC: formal analysis, visualization. XEC: investigation. WHZ: methodology, formal analysis. JK: investigation. RH: investigation. YL: investigation. HSY: resources. YX: investigation. JSH: conceptualization, resources. SPH: investigation. YL: investigation. HL: conceptualization. LZ: conceptualization. MXJ: investigation. JL: resources. KMK: conceptualization, formal analysis, resources, writing - review & editing, visualization, supervision, project administration, funding acquisition. RZ: conceptualization, resources, writing - review & editing, visualization, supervision, project administration, funding acquisition.

## Data sharing

Individual participant data and the data dictionary defining each field in the set will not be made available as all participants did not consent to have their data as a public resource. The group-level data results as well as the data processing code which do not disclose the participants’ information will be available with publication from the corresponding authors (KMK and RZ) on reasonable request (including a research proposal), subject to review.

## Acknowledgments

This work was supported by Key Technological Projects of Guangdong Province (grant number 2018B030335001 - KMK), Beijing Municipal Science & Technology Commission (grant number Z181100001518005 - RZ), the National Key Research and Development Program of China (2016YFC0105501 - RZ), Sichuan Science and Technology Program (2021YJ0247 - XJ), National Natural Science Foundation of China (61976045 - XJ, 81801779 - SXJ). The funders played no role in the writing of the manuscript or the decision where to submit it for publication. Furthermore, the authors would like to thank Dr. Ruo-Yan Zhao for her GDS assessment. We appreciate Ms. Qing-Yun Wei of Sunshine Friendship Rehabilitation Center, Beijing, China, and Ms. Meng-Lin Sun of Wucailu Rehabilitation Center, Beijing, China, for their cooperation of children recruitment. Finally, we are grateful to all the participant children and their families.

