## Supplemental Material for "A brain structural connectivity biomarker for diagnosis of autism spectrum disorder in early childhood"

**SUPPLEMENTARY MATERIAL:**

**SUPPLEMENTARY METHODS**

**MRI Acquisition and Preprocessing**

For ASD validation dataset MRI images were acquired on a GE 3T MR750 scanner with an 8-channel head coil at the University of Electronic Science and Technology of China. DTI data were acquired with an echo-planar imaging sequence: TR = 8,500 ms, FOV = 256 mm, matrix size = 128 × 128, voxel size = 2 mm isotropic, 60 slices covering the whole brain with no gap, 32 diffusion directions, and b-value = 1000 s/mm^2^.

For TD validation dataset MRI images were acquired on a 3T Verio MRI system (Siemens Medical Systems, Germany) with a birdcage gradient head coil at Nanjing Brain Hospital. DTI data were scanned with a single-shot echo-planar sequence: TR = 9,000 ms, TE = 104 ms, flip angle = 90°, FOV = 230 mm, matrix size = 128 × 128, voxel size = 1·8 mm × 1·8 mm × 2·5 mm, 60 slices covering the whole brain with no gap, NEX = 2·0, 32 diffusion directions, and b-value = 1000 s/mm².

### Pre-processing for DTI included skull removal, motion correction, eddy correction, and tensor fitting via FSL.^1^ Specifically, the most widely used FA (fractional anisotropy) measure in DTI was adopted in this study. Deterministic streamline fibers were then reconstructed from the pre-processed DTI based on the diffusion tensor model via DSI Studio.^2^ The major fiber tracking parameters were the same as in previous studies^3^: fiber count = 40,000, step size =1 mm, max turning angle = 60°, minimum fiber length = 30 mm, maximum fiber length = 300 mm, smoothing = 1.

**Two Approaches for Alleviating the Potential Effect of Imbalanced Sample Size between ASD and TD**

The first approach was the well-known SMOTE (Synthetic Minority Oversampling Technique algorithm) which upsampled the minority group of data samples by taking random simple replication of nearest neighbour samples to achieve balance between the two groups.^4^ In this study, we upsampled the TD sample size to 93 to match the ASD sample size. The second approach was down-sampling. Each time we randomly selected 26 out of 93 samples from the ASD group to train the classification model together with all 26 TD samples, and repeated the procedure 1,000 times to obtain the averaged classification accuracy.
